# State-level trends in urban-rural differences in cigarette smoking in the United States

**DOI:** 10.64898/2026.08.06.26359850

**Authors:** Clayton Ulm, Shelley D. Golden, Frantasia Hill, Christopher A. Wiesen, Sarah D. Mills

**Author notes:** Corresponding author (CU).

## Abstract

**Introduction:** Smoking prevalence remains higher in rural than in urban populations in the United States. To examine recent trends, we assessed state-level differences in cigarette smoking between urban and rural areas from 2018 to 2024.

**Methods:** Using repeated cross-sectional data from the Behavioral Risk Factor Surveillance System, we estimated state-specific logistic regression models to examine the relationship between urban-rural county residence and cigarette smoking. Unadjusted models (model 1) included urban-rural county status and year. Subsequent models (model 2) added age, sex, and race/ethnicity. A final model (model 3) included education and an interaction term between urban-rural county status and year to examine whether gaps in urban-rural smoking changed over time. In states with significant interactions, simple effects tests compared trends for urban-rural groups separately.

**Results:** Compared to urban adults, rural adults had higher unadjusted odds of cigarette smoking (odds ratio [OR] range:1.07-1.88) in 88.4% (38/43) of states. Adjusting for demographic covariates (model 2) increased the proportion of states with significant marginal effects of rurality to 90.7% (ORs:1.09-1.87). A final model that also controlled for education (model 3) decreased the proportion of states with significant marginal effects of rurality to 60.5% (ORs:1.10-1.54). Among the 14 states with significant interaction terms, the odds of smoking declined faster among urban than rural residents.

**Conclusion:** Urban-rural differences in smoking persist across most states. No state showed a reduction in urban-rural disparities over time, and the urban-rural gap widened in 14 states. Demographic variation accounted for some, but not the majority, of observed urban-rural differences.

## INTRODUCTION

Overall, smoking prevalence has markedly declined in the United States (US) for several decades. However, smoking prevalence remains higher in rural as compared to urban populations, and this difference has grown over time.[1,2] In 2020, smoking prevalence was 4.8 percentage points higher among individuals living in rural as compared to urban counties (19.2% vs 14.4%) in the US.[1] The urban-rural difference in smoking starts during adolescence, as rural middle and high school students are 43% and 64% more likely to smoke, respectively, compared to urban students.[3,4] Rural adolescents are also more likely to report exposure to secondhand smoke at home and in the car.[5] Rural differences in cigarette smoking contribute to a higher burden of smoking-related diseases in rural areas. Rural populations have nearly double the incidence of developing lung cancer compared to urban populations, and this difference is primarily attributed to higher smoking prevalence rates in rural populations.[6–8]

Rural areas differ from urban areas in terms of the composition of smoking-related demographic characteristics. Rural residents tend to have lower educational attainment and are more likely to be White, and older than urban residents.[9] However, research suggests that these factors do not fully explain the urban-rural difference in smoking prevalence in recent years.[2] Using nationally representative data from the National Survey on Drug Use and Health, Doogan et al.[2] found that demographic (age, sex, race/ethnicity, education, income, marital status, employment status, and type of occupation) and psychosocial covariates (anxiety, depression, substance abuse) fully explained urban-rural differences in smoking in 2007-2008. However, these covariates failed to explain the widening difference in urban-rural smoking by 2014-2016.[2,3] This suggests that, at the national level, other factors may be associated with lower smoking rates in urban than in rural areas.[2,3] If state-specific analyses reveal that the widening difference observed at the national level persists across all states, this may suggest a universal explanation. However, if it is more pronounced in some states, this may indicate the need to explore state-specific policies and contexts as critical contributors to the difference.

To understand recent trends in urban-rural differences in smoking, this study: 1) provides smoking prevalence estimates in urban and rural counties in each US state in 2018 and 2024, 2) examines whether demographic variations in urban and rural counties account for urban-rural differences in the odds of smoking, and 3) assesses whether urban-rural differences in smoking have widened, narrowed, or experienced no change from 2018 to 2024 in each state. Analyses were conducted within each state to determine whether trends are shared across different state contexts. Findings from this study may help identify which states may benefit from targeted tobacco control intervention as well as state-specific contexts that either sustain or exacerbate urban-rural differences in smoking.

## METHODS

This is a secondary analysis of repeated cross-sectional data from the BRFSS, an annual state-representative survey of US adults that collects information on health behaviors, including tobacco use.[10] The BRFSS is a landline and cellular phone survey sponsored by the Centers for Disease Control and Prevention (CDC).[10] It is administered to more than 400,000 respondents across all 50 states, Washington, D.C., and the US territories. In this study, we used 2018 to 2024 data from the BRFSS Core Survey.

### Sample

Using BRFSS data, we created two separate datasets to examine trends in current cigarette smoking by urban-rural county status at the national level and among US states. For the nationwide analysis, we removed individuals with missing values on any demographic characteristics included in the regression models. From an initial dataset of 3,032,480 respondents, 2,741,600 (90.4%) individuals were included after excluding those with missing demographic information. For the state-specific analyses, we also removed eight states (Connecticut, Delaware, District of Columbia, Hawaii, Massachusetts, New Hampshire, New Jersey, and Rhode Island) that did not contain any BRFSS respondents living in rural counties, leaving 43 states comprising 2,417,724 total respondents with complete demographic data for state-specific analyses.

This study was reviewed by the University of North Carolina at Chapel Hill Office of Human Research Ethics (study #24-0758), which determined that this secondary analysis of publicly available, de-identified data does not constitute human subjects research as defined under federal regulations (45 CFR 46.102 and 21 CFR 56.102) and does not require institutional review board approval. Informed consent was not applicable because the study did not involve interaction with human participants.

### Measures

#### Current Smoking

The BRFSS Core Survey asks participants if they have smoked at least 100 cigarettes in their lifetime and if they currently smoke cigarettes “every day,” “some days,” or “not at all.” We defined current smokers as those who reported smoking at least 100 cigarettes in their lifetime and currently smoke “every day” or “some days.”

#### Demographic Characteristics

The BRFSS collects the following demographic information from individual participants: age, sex (male or female), race/ethnicity (non-Hispanic White, non-Hispanic Black, non-Hispanic Other race/Multiracial, and Hispanic), education level (did not graduate high school, graduated high school, attended college or technical school, and graduated from college or technical school), state, survey year (2018, 2019, 2020, 2021, 2022, 2023, 2024), and urban-rural county status. The BRFSS classifies the county of each participant as urban or rural using the 2013 National Center for Health Statistics (NCHS) Urban-Rural Classification Scheme for Counties, derived from the Office of Management and Budget’s 2013 classification of metro and micropolitan statistical areas.[11] Participants with counties in the NCHS categories 1 through 5 (large central metro, large fringe metro, medium metro, small metro, micropolitan) are classified as urban; otherwise, they are in NCHS category 6 (non-core) and are considered rural.[11]

### Data Analysis

To describe sample characteristics, we calculated weighted descriptive statistics for aggregated survey data (2018-2024). In 2018 and 2024, we also calculated weighted cigarette smoking prevalence estimates by state and urban-rural county status in each year. To follow CDC guidelines about reporting on and interpreting BRFSS data, in each year, we assessed the relative standard error (RSE) of cigarette smoking prevalence estimates for each urban-rural category based on the unweighted sample size for each subgroup in each state.[12] If the RSE was greater than 30% or the unweighted sample size was less than 50, the smoking prevalence estimate for that state in that year was excluded to prevent misinterpretation of unreliable estimates.[12]

In nationwide and state-specific analyses, we used a set of four logistic regression models to examine associations of urban-rural county status and current cigarette smoking. **Model 1** included only urban-rural and year as predictor variables. This model provided the unadjusted odds estimates for urban-rural and served as a baseline for comparison with subsequent models. We progressively added demographic covariates to Models 2 and 3 to assess their impact on urban-rural differences in smoking. **Model 2** added age, sex, and race/ethnicity covariates to evaluate the marginal effect of rurality after controlling for these demographic factors. To also examine the impact of educational differences in urban-rural populations, **Model 3a** also included education as a covariate. Finally, to determine whether the odds of smoking in urban and rural areas have changed over time, an interaction term between year and urban-rural status was added in **Model 3b**. For states with significant interaction terms, simple effects tests were conducted to compare time trends for urban and rural groups separately. We estimated regression models and conducted simple effects tests using SAS 9.4 (SAS Institute Inc.) and the BRFSS complex survey design procedures. We generated survey-weighted predicted probability plots in Stata SE 19 (StataCorp LLC) and produced maps in Python using geopandas and matplotlib.[13,14]

## RESULTS

Table 1 provides demographic characteristics of the study sample from 2018 to 2024. In the national sample, the prevalence of cigarette smoking declined each year over this period. From 2018 to 2024, smoking prevalence estimates were highest among rural residents (19.0%), individuals 55-64 years (16.3%), non-Hispanic Multiracial individuals (19.3%), and those that did not graduate high school (19.2%). Among rural counties, state-level smoking prevalence estimates ranged from 10.8% (Washington) to 28.7% (Tennessee) in 2018 and 8.6% (Arizona) to 22.0% (West Virginia) in 2024. Among urban counties, smoking prevalence estimates ranged from 8.6% (Utah) to 24.2% (West Virginia) in 2018 and from 5.3% (Utah) to 19.3% (West Virginia) in 2024 (see S2 Table and S1 Fig).

**Table 1.** Sample demographic characteristics (N = 1,799,666,601), BRFSS 2018-2024.

| Variable | Sample size | Percentage | Smoking prevalence |
| --- | --- | --- | --- |
| **County** |  |  |  |
| Urban | 1,663,625,431 | 92.4% | 13.0% |
| Rural | 112,737,158 | 6.3% | 19.0% |
| Missing | 23,304,012 | 1.3% | 9.6% |
| **Age in years** |  |  |  |
| 18-24 | 218,854,389 | 12.2% | 7.7% |
| 25-34 | 308,254,641 | 17.1% | 15.1% |
| 35-44 | 295,808,375 | 16.4% | 17.1% |
| 45-54 | 282,448,503 | 15.7% | 15.7% |
| 55-64 | 291,969,733 | 16.2% | 16.3% |
| 65 or older | 402,330,960 | 22.4% | 8.7% |
| **Sex** |  |  |  |
| Female | 922,308,313 | 51.2% | 11.7% |
| Male | 876,738,603 | 48.7% | 15.1% |
| Missing | 619,685 | <0.1% | 13.3% |
| **Race/ethnicity** |  |  |  |
| Non-Hispanic White | 1,053,914,781 | 58.6% | 14.0% |
| Non-Hispanic Black | 208,365,494 | 11.6% | 15.0% |
| Non-Hispanic Other | 144,250,163 | 8.0% | 10.3% |
| Non-Hispanic Multiracial | 28,217,309 | 1.6% | 19.3% |
| Hispanic | 323,321,601 | 18.0% | 10.8% |
| Missing | 41,597,253 | 2.3% | 13.7% |
| **Education** |  |  |  |
| Did not graduate high school | 362,364,884 | 20.1% | 19.2% |
| Graduated high school | 503,636,786 | 28.0% | 15.8% |
| Attended college or technical school | 467,534,127 | 26.0% | 14.1% |
| Graduated from college or technical school | 456,401,264 | 25.4% | 5.5% |
| Missing | 9,729,540 | 0.5% | 9.2% |
| **Survey year** |  |  |  |
| 2018 | 258,073,387 | 14.3% | 15.5% |
| 2019 | 252,430,291 | 14.0% | 15.3% |
| 2020 | 260,408,470 | 14.5% | 14.2% |
| 2021 | 246,041,640 | 13.7% | 13.4% |
| 2022 | 264,789,594 | 14.7% | 12.8% |
| 2023 | 254,139,829 | 14.1% | 11.4% |
| 2024 | 263,783,390 | 14.7% | 10.8% |

Note. Values are weighted to represent the U.S. adult population. The unweighted sample sizes across states ranged from 19,720 (Nevada) to 182,925 (New York) respondents. Within states, the rural sample sizes ranged from 580 (Nevada) to 42,538 (Maine) respondents, and urban sample sizes ranged from 19,140 (Nevada) to 167,203 (New York) respondents. “Smoking prevalence” is the percentage of current cigarette smokers within the total sample for each subgroup. For example, 6.3% of the total weighted sample lived in rural counties, and 19.0% of adults in those counties were current smokers.

### Current Smoking

Odds ratios of urban-rural differences in current cigarette smoking for each state are presented in Fig 1 and S1 Table. In the nationwide analysis, rural residents had significantly higher odds of cigarette smoking in Model 1 (OR = 1.57, 95% CI: 1.43, 1.71), Model 2 (OR = 1.55, 95% CI: 1.44, 1.67), and Model 3a (OR = 1.31, 95% CI: 1.22, 1.41). In state-specific analyses, in Model 1 (unadjusted) there were significant marginal effects of rurality in 90.7% (39/43) of states, with rural residents having higher odds of cigarette smoking than urban residents in 38 states (OR range: 1.07-1.88). Adjusting for demographic covariates (age, sex, and race/ethnicity) in Model 2 maintained this proportion, with 90.7% (39/43) of states showing higher odds of smoking among rural residents (OR range: 1.09-1.87). After also including education as a covariate in Model 3a, the proportion of states with significant estimates decreased to 62.8% (27/43). Rural residents had higher odds of current smoking in 26 of these states (OR range: 1.10-1.54). In the other 16 states, there was no significant difference (P > .05) in the odds of smoking among urban and rural residents. In Wyoming, rural residents had lower odds of smoking (OR = 0.89, 95% CI: 0.81, 0.99) than urban residents.

**Fig 1.**
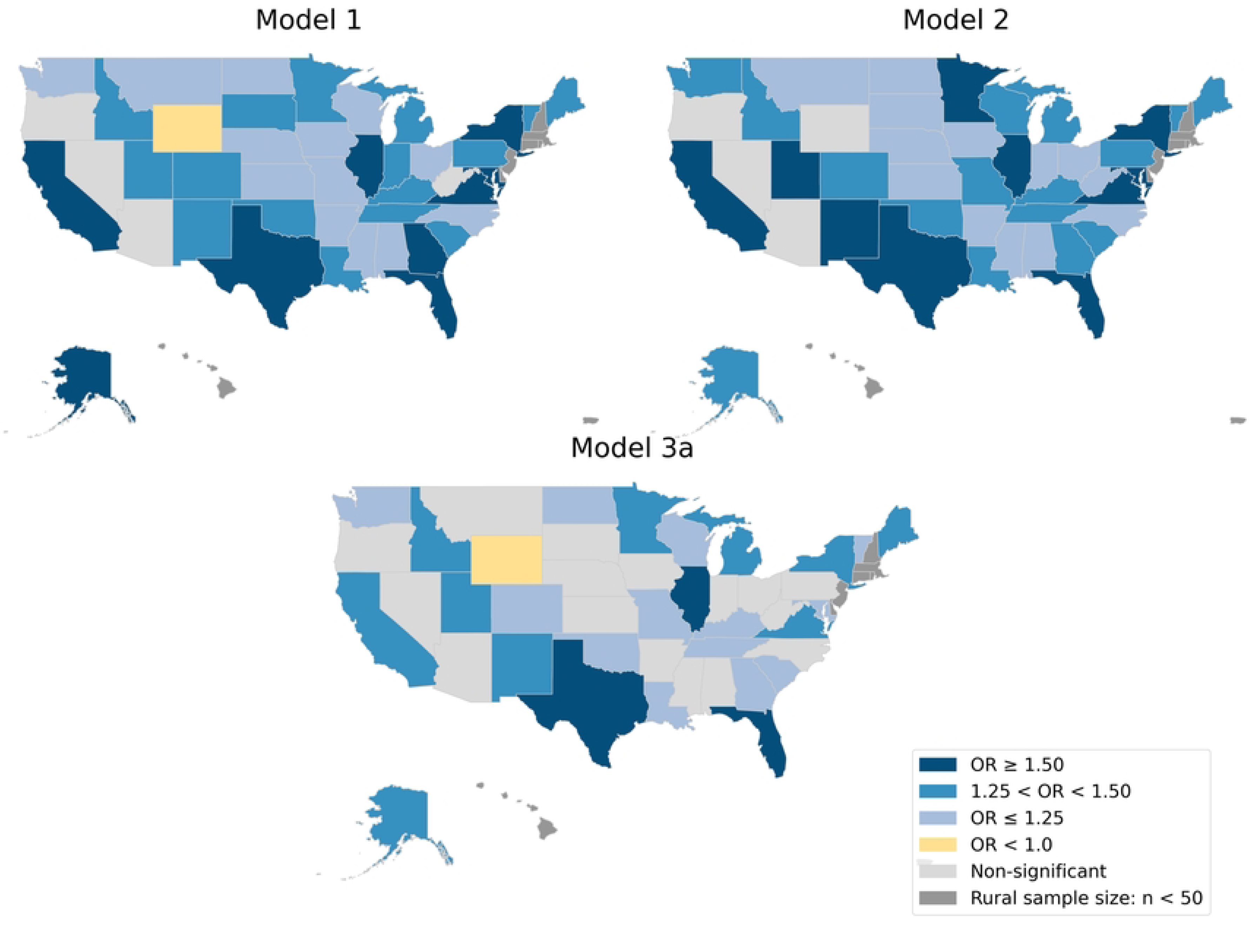
Differences in urban-rural cigarette smoking in US states, 2018-2024. Notes. OR = Odds Ratio. Odds of smoking were higher among rural residents in darker-colored states. States with non-significant rural to urban odds ratios or insufficient rural sample sizes (n < 50) for analysis are in light and dark grey, respectively.

### Interaction Effects of Urban-Rural and Year

To examine whether the odds of smoking in urban and rural areas changed over time, we included an interaction term between urban-rural and year in Model 3b. Nationwide, we found that urban residents had faster declines in current smoking than rural residents from 2018 to 2024 (P for interaction in Model 3b < .001). In state-specific analyses, the interaction term was not significant in 29 states, indicating no difference in the rate of decline in the odds of smoking between residents of urban and rural counties from 2018 to 2024 in these states. However, there were 14 states with significant interaction terms.

Table 2 presents the results of simple effects tests among the 14 states with significant interaction terms. The odds ratios from the simple effects tests represent the annual change in the odds of current smoking over time in urban and rural groups separately. In each of the 14 states (Arizona, Arkansas, Colorado, Georgia, Iowa, Kansas, Maine, Mississippi, Missouri, Montana, Nebraska, South Carolina, Washington, and Wisconsin), faster declines in the odds of smoking were observed among urban residents (all P values < .05). For example, in Arizona the odds of smoking significantly decreased by 7% each year among urban residents (OR = 0.93, 95% CI: 0.90, 0.97) from 2018 to 2024, but there was no significant change in the odds of smoking among rural residents (OR = 0.98, 95% CI: 0.90, 1.06). In Wisconsin, the odds of smoking decreased by 9% each year among urban residents (OR = 0.91, 95% CI: 0.89, 0.94), but by only 6% each year among rural residents (OR = 0.94, 95% CI: 0.90, 0.98). Extracted from Model 3b, Fig 2 visualizes the predicted probabilities of smoking among urban and rural groups each year from 2018 to 2024 in states with significant interaction terms.

**Fig 2.**
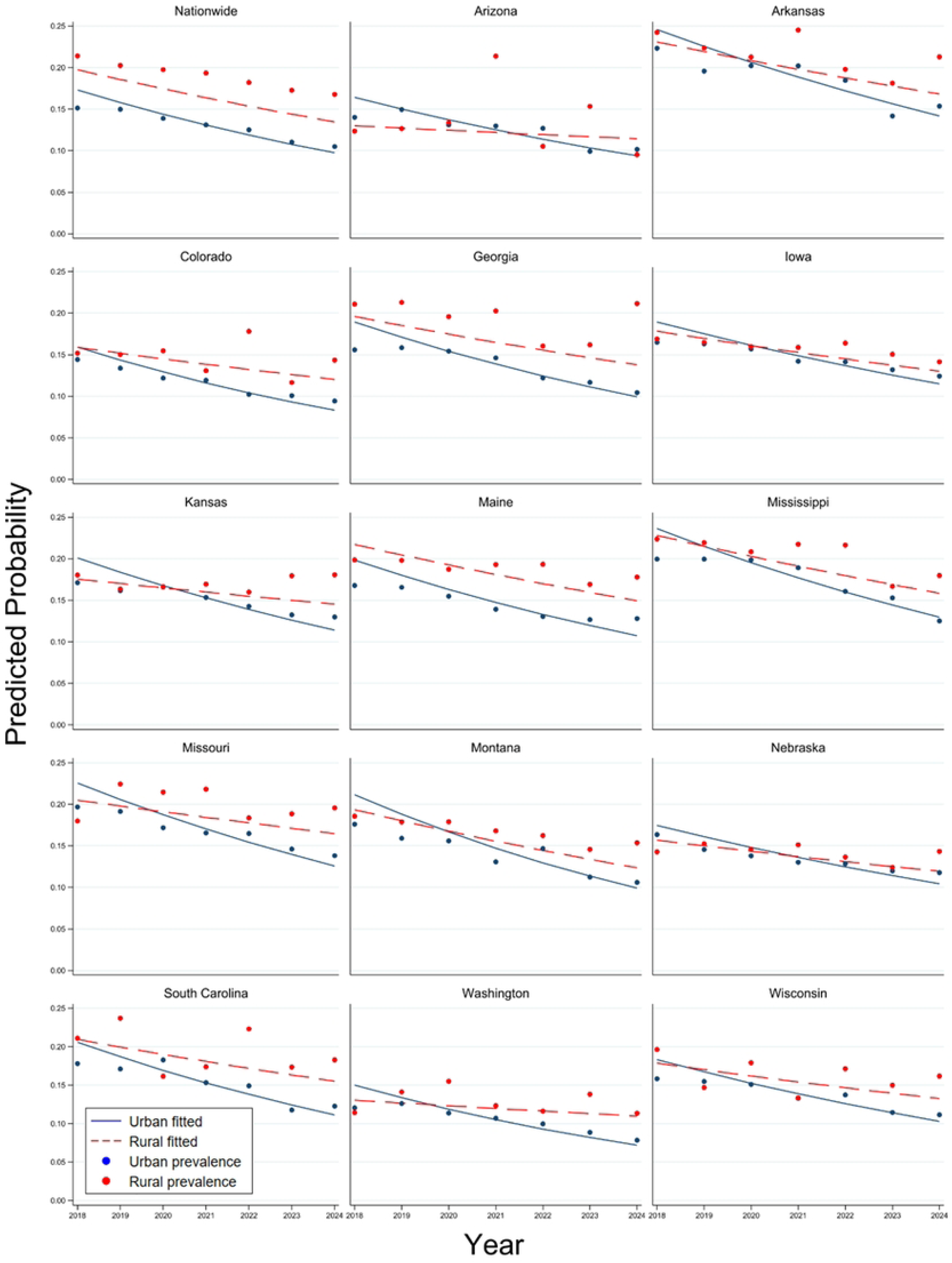
Predicted probabilities (lines) from logistic regression Model 3b and unadjusted prevalences (dots) of current smoking in rural (red) and urban (blue) counties nationwide and in states with significant interaction terms, 2018-2024.

**Table 2.** Linear time trends in current smoking among urban and rural groups.

|  | Urban |  | Rural |  |
| --- | --- | --- | --- | --- |
|  | OR | 95% CI | OR | 95% CI |
| Nationwide | 0.91 | (0.90, 0.91) | 0.92 | (0.91, 0.93) |
| Arizona | 0.93 | (0.90, 0.97) | 0.98 | (0.90, 1.06) |
| Arkansas | 0.91 | (0.89, 0.93) | 0.93 | (0.90, 0.97) |
| Colorado | 0.91 | (0.89, 0.94) | 0.95 | (0.90, 1.00) |
| Georgia | 0.90 | (0.88, 0.93) | 0.93 | (0.88, 0.98) |
| Iowa | 0.92 | (0.90, 0.93) | 0.94 | (0.91, 0.96) |
| Kansas | 0.92 | (0.91, 0.94) | 0.96 | (0.93, 0.99) |
| Maine | 0.90 | (0.88, 0.91) | 0.92 | (0.90, 0.94) |
| Mississippi | 0.90 | (0.88, 0.92) | 0.92 | (0.89, 0.96) |
| Missouri | 0.92 | (0.90, 0.94) | 0.95 | (0.92, 0.99) |
| Montana | 0.88 | (0.86, 0.90) | 0.91 | (0.88, 0.94) |
| Nebraska | 0.92 | (0.91, 0.94) | 0.95 | (0.93, 0.97) |
| South Carolina | 0.91 | (0.88, 0.94) | 0.94 | (0.88, 0.99) |
| Washington | 0.92 | (0.89, 0.94) | 0.97 | (0.92, 1.02) |
| Wisconsin | 0.91 | (0.89, 0.94) | 0.94 | (0.90, 0.98) |

Note. OR = Odds Ratio, CI = Confidence Interval. Example interpretation: In Arizona, the odds of smoking decreased by 7% each year (OR = 0.93, 95% CI: 0.90, 0.97) among urban residents, but did not decrease significantly (OR = 0.98, 95% CI: 0.90, 1.06) for rural residents.

## DISCUSSION

From 2018 to 2024, cigarette smoking prevalence in rural counties remained higher than in urban counties across most US states. In 2024, the greatest urban-rural differences in unadjusted cigarette smoking prevalence were in California, Idaho, Illinois, Georgia, and Virginia, where smoking prevalence estimates were twice as high among residents of rural as compared to urban areas. Even after adjusting for demographic characteristics, including educational differences, individuals living in rural counties had significantly higher odds of smoking in more than half (60.5%) of states. Findings from this study suggest that demographic differences in urban and rural residents account for some of the differences in smoking prevalence estimates. However, other factors, such as local and state tobacco regulatory policy environments, may also play an important role.

To our knowledge, this is the first study to assess whether urban-rural differences in cigarette smoking have widened, narrowed, or remained unchanged in each US state over time. Our analyses revealed state-level heterogeneity in trends. In the majority (67%) of states, there was no change in the odds of smoking between urban and rural areas over time. In 14 states, however, we found that the urban-rural gap widened. This widening gap can be attributed to more rapid declines in the odds of smoking among residents of urban counties within these states, but relatively slower declines or stagnation among residents of rural counties. From 2018 to 2024, no state reduced urban-rural differences in the odds of smoking. Findings from this study suggest targeted tobacco control efforts in rural communities are needed to help reduce the urban-rural disparity in cigarette smoking.

### Policy Implications

One potential explanation for the persistent urban-rural gap in smoking prevalence is the unequal implementation of tobacco control policies in rural areas. Research indicates that people living in rural areas are often less protected by evidence-based tobacco control policies.[15–17] For example, compared to suburban or urban areas, rural communities have been slower to implement smoke-free parks and sales restrictions on flavored tobacco products.[15,18] In Texas, which had one of the greatest urban-rural smoking differences in our analysis, only 3-5% of localities in the state had comprehensive smoke-free air laws in 2011, and almost all of these were in urban areas.[17] As of 2025, Texas still has no statewide comprehensive smoke-free law, leaving many rural areas unprotected.[19] Weaker smoke-free air laws can reinforce positive social norms around smoking.[20] Worksites in rural counties are also less likely to have tobacco-free policies than those in urban counties.[21] In addition, enforcement of tobacco control policies has typically been higher in urban areas, with rural areas overrepresented among those with no tobacco retailer inspections.[22]

Research suggests that the rural-urban disparity in the adoption and enforcement of tobacco control policies is influenced by factors such as political resistance, limited public health resources, and less developed public health infrastructure in rural areas.[15,18] Strengthening state-level tobacco control efforts (for example, raising state tobacco taxes or enacting comprehensive smoke-free air laws that cover all workplaces and venues) can protect rural residents with more limited local tobacco control measures. Implementing tobacco control measures that are more effective in rural (as compared to urban) areas, such as population-based retailer licensing caps (e.g., 1 per 1,000 residents) rather than proximity-based caps (e.g., within 500 feet of other retailers),[23] could further reduce urban-rural differences in tobacco retailer density, which studies show is associated with tobacco use.[24] Enabling rural municipalities to implement and enforce policies by removing legal barriers, such as preemption, could also address the persistent urban-rural gap in cigarette smoking.

### Strengths and Limitations

This study has notable strengths. We conducted state-specific analyses using a large, state-representative survey over multiple years. We also estimated a series of regression models with different demographic covariates in an effort to isolate the marginal effect of rurality on current cigarette smoking. Study limitations should also be considered. We did not include all potential covariates that may influence smoking behavior, such as income, healthcare access, and mental health. These variables might also explain observed urban-rural differences. In addition, urban-rural was dichotomized in this study, which does not reflect the spectrum of rurality. Also, we only included a linear time trend in the regression models and did not consider potential non-linear time trends. A preliminary assessment of the quadratic time trend, however, indicated it was not significant in most states. Finally, tobacco control and other policy variables were not examined in this study. Future research should examine how local and state policies are associated with urban-rural differences in cigarette smoking to identify potential pro-equity tobacco control policy interventions.

### Conclusions

Findings from this study suggest that living in rural areas presents a unique vulnerability to cigarette smoking. While demographic differences in urban and rural populations account for part of the increased risk, they do not fully explain it. In addition, from 2018 to 2024, no US states reduced the urban-rural gap in cigarette smoking. Findings from this study help to identify US states that are a priority for tobacco control intervention to reduce urban-rural disparities in cigarette smoking.

## Data Availability

Data used in this secondary data analysis are publicly available from the CDC Behavioral Risk Factor Surveillance System (https://www.cdc.gov/brfss/annual_data/annual_data.htm). Code used for this project is listed on a publicly available GitHub repository (https://github.com/rulm0000/Smoking-Prevalence-Analysis-2018-2024).

## Supporting information

**S1 Fig.** State-level smoking prevalence estimates in rural and urban counties. The maps show smoking prevalence estimates for 2018 (left panels) and 2024 (right panels) in rural (top panels) and urban (bottom panels) counties. Higher prevalence is indicated by red and lower prevalence by green. State estimates are excluded (grey) if the unweighted sample size was less than 50 or the relative standard error was greater than 30%.

**S1 Table.** Odds ratios for the urban-rural variable in the nationwide and state-specific logistic regression models.

**S2 Table.** State-level smoking prevalence estimates in rural and urban counties, 2018 and 2024. Values are blank if the relative standard error was greater than 30% or the unweighted sample size was less than 50.

**S1 Checklist.** STROBE checklist for cross-sectional studies.

